# Understanding the Intersection between Midwives’ Culture, Educational Background and Community Practice in Neonatal Jaundice Care in Ghana: A Qualitative Inquiry

**DOI:** 10.64898/2026.04.18.26350907

**Authors:** Gloria Asamoah, Mary Ani-Amponsah, Caroline Dinam Badzi

## Abstract

Culture plays a crucial role in health; family, community, culture, and social conventions all have a significant impact on how an infant with jaundice is treated. Written or unwritten rules govern what parents and the community are allowed to do, which may have a detrimental effect on the neonate’s care.

**Objectives:** The study explored how social expectations affect midwives’ management of neonatal jaundice at the St Patrick’s hospital in Maase-Offinso, in the Ashanti region of Ghana.

**Method:** A total of seventeen midwives were sampled purposively using an exploratory descriptive design. Participants were engaged in interviews and focus group discussion after ethical approval was obtained. A semi-structured focus group discussion guide and interview guide was used to collect data.

**Results:** The study discovered that the treatment of neonatal jaundice was adversely affected by social pressures, misconceptions, maternal choices, and spiritual views. Mothers and midwives socially approved sunbathing, and there were indications that grandmothers disapproved hospital care for their grandchildren.

**Conclusion:** Culture, family and social norms cannot be separated from health especially for the neonate whose means of identification is to belong to a family. Consequently, it is essential to respond to social influences, cultural conventions, and the various cultures of families with a culturally sensitive approach.

## 1.0 Introduction

While previous research frequently ignored the contribution of culture to economic growth (Guiso et al., 2006) and even discovered that socioeconomic variables, rather than sociocultural practices, are directly linked to infant mortality (Gyimah, 2006). There is compelling evidence that Ghanaian cultural customs have an impact on children’s health (Nott, 2018). Experts in modern development recognize its significance (Arestis et al., 2021). They are aware that viewpoints, values, customs, and beliefs are essential for long-term socioeconomic advancement (Zheng et al., 2021). In Ghana, people’s attitudes and behaviors are greatly influenced by cultural beliefs, social standards, and spirituality, particularly when it comes to health and the selection of medical treatments. Despite modernization, traditional cultural methods are still used in African health care today (Ibrahim & Ahmedolaitan, 2022). Infant morbidity and mortality in Ghana have been partially caused by cultural practices and attitudes about child health care (Sumankuuro et al., 2019).

Traditional medicine, which is frequently firmly anchored in cultural customs, provides all-encompassing methods that connect with local populations and attend to both physical and spiritual health issues (Kendi, 2024). Traditional medicine, which is frequently firmly anchored in cultural customs, provides all-encompassing methods that connect with local populations and attend to both physical and spiritual health issues. Therefore, understanding the culture in the nurse’s service area, particularly with regard to healthcare practices and beliefs, will help the nurse avoid making biased assumptions about the client’s culture and become conscious of his own biases and prejudices in order to plan and provide culturally competent care (Maniago, 2020). The norms of the society to which a newborn and family belong have a significant impact on their health, particularly in a multicultural nation like Ghana. Social norms are significant determinants of health-related behaviors and are included in several well-known psychological theoretical models of health behavior (Ajzen, 1991).

Nearly every ailment that affects a newborn in Ghanaian communities is referred to as “Asram”; some claim it is a conventionally defined illness, while others claim it is spiritual. A number of childhood illnesses are still considered spiritual in Ghana and are described elsewhere as quite different. Previous research has characterized Asram as potentially severe and as a spiritual illness that can be transmitted from a pregnant woman to an unborn child, despite the fact that Asram does not appear to have a biological equivalent (Meñaca et al., 2017). According to (Acheampong et al., 2022), Asram was classified based on the signs the sick child presented Asram Boredwo (a wasted child with a larger head and drier skin), Asram ntoos (a description of several blisters on a baby’s skin), Asram Bofre (a swollen and larger-than-normal head), Asram mpompo (a baby with rashes all over the body), Asram mpaemu (a sign that the child’s skull has split into two or four), and Asram ayamtuo (children with a bloated stomach and typically have watery stools during the first week of life), Asram nofo-denden, which causes a mother’s breast to become hard and hot with a hardened nipple; Asram esuro, which causes a sudden spike in temperature and convulsions with clenched fingers and wide-open eyes between the ages of one and five; and Asram pepe, which causes rapid breathing and is thought to be the most dangerous because it frequently results in death.

Rules and standards that are understood by members of a group, and that guide or constrain social behaviors without the force of law” is the conventional definition of social norms. According to Cialdini and Trost, (1998), they frequently have to do with perceived social pressure to participate in or refrain from particular activities (Ajzen, 1991).

The Social Norm Approach (SNA) has its origins in a study conducted by (Perkins & Berkowitz, 1986). According Cialdini et al., (1991) social norms, which may be quantified in connection to various social networks, comprise both descriptive and injunctive norms that have an impact on behavior. Perceptions of a social network’s behavior, such as that of classmates, family, and adults, are known as descriptive norms. Perceptions about what members of a social network should or shouldn’t do are known as injunctive norms. Also known as “behavioral norms,” descriptive norms show which behaviors are common (Perkins, 2002). Other theory mentioned normative social behavior, example Rimal and Real, (2005) describes and forecasts the effects of social norms on behaviors. Therefore, normative influence, which is the effect of social norms on individual behavior, indicates that the environment a midwife finds herself in along with her personal norms cannot be undermined. These constructs interact because midwives may be more likely to engage in a behavior if they believe it to be both common (descriptive norm) and socially acceptable (injunctive norm). The impact of these social norms on midwifery practice (normative influence) is largely determined by groups that individuals identify with and whose norms they follow.

Being conscious of one’s own attitudes and dispositions toward other ethnic groups enables midwives to give holistic care and adapt to sociocultural norms affecting their care supply and management for newborn jaundice (Albougami et al., 2019). Family values and the primary caregiver’s spiritual or religious views play a crucial role in influencing the decision-making and healthcare-seeking behaviors that impact the newborn’s care.

Research on social expectations in health settings has concentrated on the health-seeking behaviors of primary caregivers, but it has not examined how these norms affect the way care professionals provide care. For the midwife, normative expectations, psychological well-being derived from conformity, avoiding actions they believe will be disapproved to maintain their social standing, and finally, their personal norms are crucial in decision-making (Gavrilets et al., 2024). The spiritual belief, perceived misconceptions, what is approved or disapproved in the midwife’s environment can affect their care provision.

Therefore, it is equally crucial to examine how established conventions influence midwives’ reactions and infant care, particularly when it comes to the categorization of newborn illnesses that are categorized as traditional or spiritual ailments in the Ghanaian society. This makes it ideal for the researcher, a midwife and a NICU nurse; to investigate how culture, community, and societal expectations influence midwives’ reactions to neonatal jaundice, the most common condition during the neonatal period in a multi-cultural setting like Offinso South municipality in Ghana.

This study explored how social expectations thus descriptive standards, injunctive norms, and normative influence affect midwives’ management of neonatal jaundice at the St Patrick’s hospital in Maase-Offinso, in the Ashanti region of Ghana.

## 2.0. METHOD

### 2.1. Study Design

An exploratory descriptive qualitative design was used in this study to explore how social norms affect the readiness and management of NNJ of midwives in Ghana.

### 2.2. Study Setting and Participants

A total of seventeen midwives that met the inclusion criteria were recruited purposively based on the principle of saturation. The study was conducted at the Neonatal Intensive Care Unit (NICU) and postnatal ward (PNW) of the St Patrick’s Hospital in the Offinso municipality of Ghana. To refine the researchers understanding, after the initial data gathered through the individual in-depth interviews, a focus group discussion with 2 weeks prior notice was schedule for eight midwives, 4 each from PNW and NICU. The focus group discussion (FGD) had six (6) participants instead of the eight (8) that were scheduled earlier due to an emergency at the NICU that was beyond the researchers’ control. After the FGD the final two interviews produced no new perspectives and so no more interviews were done after the seventeenth participant.

**Figure 1:**
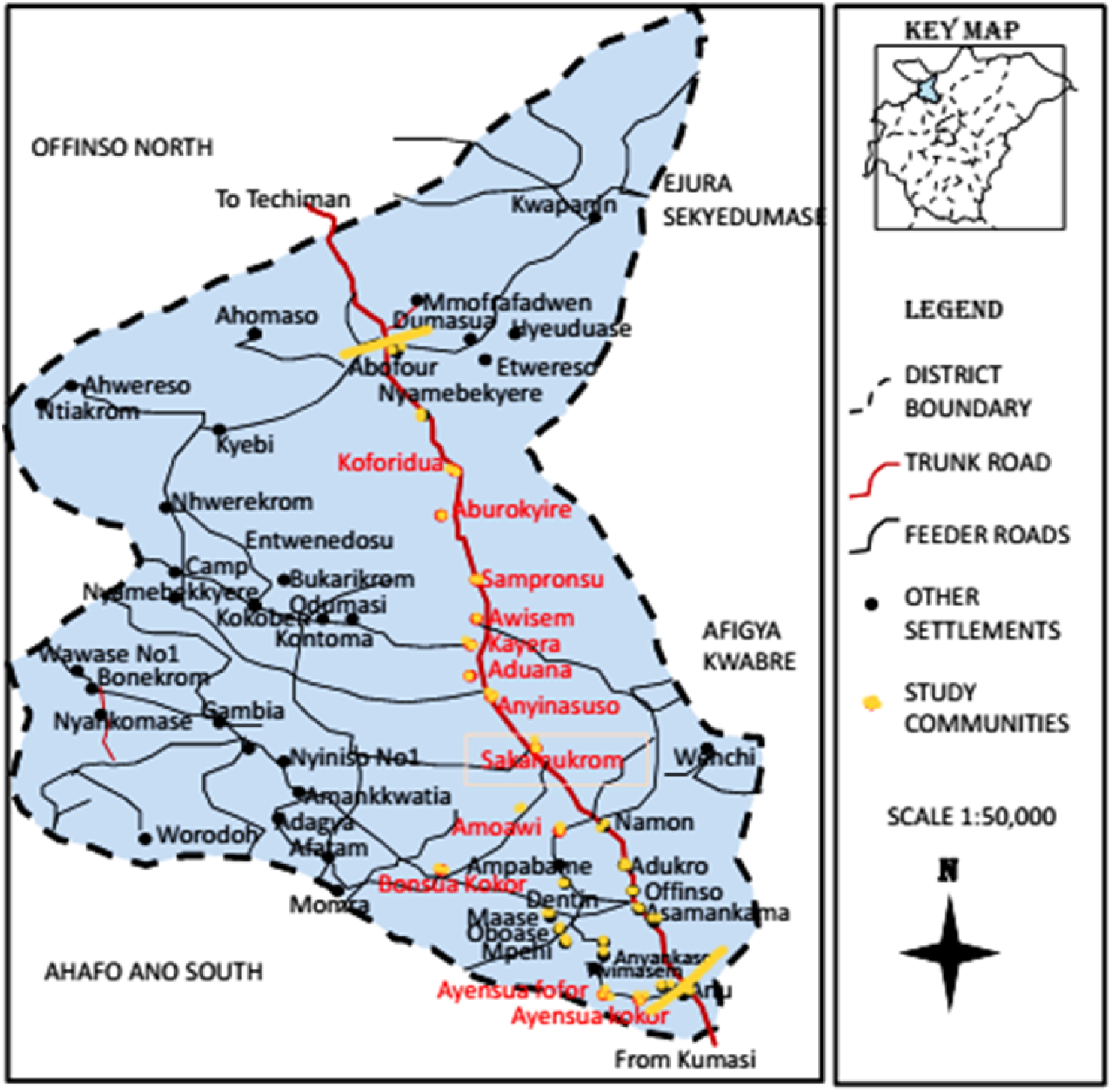
Map of Ghana showing the Offinso municipality. www.researchgate.net

### 2.3. Data Collection

Data were gathered using a semi-structured interview guide for the individual in-depth interviews and a semi-structured focus group discussion guide were used for the focus group discussion. The data collection tool was pretested using three participants before the main study.

### 2.4. Data Analysis

Data were analyzed concurrently and continuously using Braun and Clarke’s thematic analysis (Braun & Clarke, 2022). The study formed four themes descriptive norms, injunctive norms, normative influence and reference groups and eight subthemes. The study had three main themes based on constructs in the social norm; Descriptive Norms, Injunctive Norms and Normative Influence and produced nine sub themes.

### 2.5. Methodological Rigor

Lincoln and Guba, (1985) standards for assessing qualitative research were applied. According to Enworo, (2023), the study’s results and conclusions are a true depiction of the participants. For dependability, an audit trail detailing the events that occurred during the focus group discussion and interviews was maintained. In order for a study to be consistent with another study, a thorough description might help make the study applicable in a different context, making it noteworthy (Lincoln & Guba, 1985).

### 2.6. Ethical Approval

Prior to data collection, the Christian Health Association of Ghana Research Department Institutional Review Board (CHAG-IRB) granted ethics approval (CHAG-IRB05042024). Permission to perform the study at St. Patrick’s Hospital was given by the hospital administrator. Permission was also requested from the PNW and NICU unit heads.

Participants who voluntarily agreed to participate in the study without being coerced signed a consent form after receiving a thorough explanation. Participants who participated in focus group discussions were informed that privacy and confidentiality could not be maintained because everyone in the group heard what was discussed. Participants who underwent individual in-depth interviews were guaranteed confidentiality and privacy and were de-identified using an identification number.

## 3.0. Results

### 3.1. Demographic Characteristics

Eight midwives from the PNW and nine midwives from the NICU were among the seventeen midwives who took part in the study.

For participants that were engaged in the focus group discussion three participant each were from postnatal ward and NICU. All the six midwives had diploma in midwifery as their level of education for both the postnatal and NICU midwives. And the cadre of the participants were Senior Staff Midwife and Staff midwife. For the Postnatal midwives only one was a Senior Staff Midwife while two of the three NICU midwives were Senior Staff Midwife. One each of the participant from the two groups had five years of experience as midwives and the other two for PNW midwives were 1 year and 2 years while the remaining two NICU midwives had 3 years of experience. All the six participants were Christians and three were single and the other three midwives were married. Participants used were de-identified for the postnatal midwives they were assigned FP followed by the numbers 1-3 and the NICU midwives they were assigned FN followed by the numbers 1-3. The ‘F’ was the first word in focus group discussion and the ‘P’ indicated postnatal and ‘N’ also for NICU. So participants from postnatal ward had identification as FP1, FP2 and FP3 while that of NICU were FN1, FN2 and FN3.

For the eleven participants that were interviewed individually, the highest age was 43 years and the least was 25 years. Nine of the participants were aged between 25 to 35 years with two of them between 36 and 45. Most of the participants, seven out of the eleven qualification was diploma in midwifery and the remaining four had their bachelor’s degree in midwifery. The highest cadre was Senior Midwifery Officer and only a participant had that rank, five were Senior Staff Midwife, three were Midwifery Officers and two were Staff Midwife. NICU participants that were interviewed were six and five were from the postnatal ward. The highest years of experience was 12 years and the least was, the years of experience with the highest participants was five years. Only one of the participants was a Muslim, and only three of the eleven participants were single. Participants were de-identified as ID-P1 to ID-P11were ID-P was used to indicate Individual In-depth Interview Participant and the number 1 to 11 based on who was recruited first to the eleventh participant.

### 3.20. Thematic findings

Based on the data collected, three major themes descriptive standards, social expectations, and conformity pressure were developed, and the data yielded nine subthemes.

**Table 1.** Organization of Major Themes and Sub-themes.

|  | **Major themes** | **Subthemes** |
| --- | --- | --- |
| 1 | Descriptive Standards | i. Shared Beliefs about Neonatal Jaundice<br>ii. Neonatal Jaundice through a Spiritual Lens<br>iii. Following Established NICU Guidelines |
| 2 | Social Expectations | i. Grandmothers' and Families' Disapproval for Hospital Care.<br>ii. Social Approval of Sunbathing |
| 3 | Conformity Pressure | i. Midwives' Contribution to Neonatal Jaundice Care<br>ii. Protocol- informed Shared Decision-Making and Autonomy Balance<br>iii. Supervisors' Expectations: A Normative Injunctive Influence on Neonatal Jaundice Care |

### 3.21. Descriptive Standards

These speak to how common a conduct is thought to be in a community or group. People’s perceptions of NNJ and hospital treatment are described by these reported behaviors within a social group. Three subthemes formed comprised of Neonatal Jaundice Through a Spiritual Lens, Shared Beliefs about NNJ, and Following Established NICU Guidelines.

### 3.22. Shared Beliefs About Neonatal Jaundice

This subtheme had a significant impact on midwives’ treatment since the results showed that some midwives adhered to prevailing views, which influenced how they responded to and managed NNJ.

A participant mentioned when reported to a facility with her jaundice baby, the midwife asked her to expose her child under the sunlight even when she reported for hospital care:

> *“When I got to the midwife, she said three days old baby is physiological and so I should expose her under the early morning sun, but my baby was not born term so I called a doctor and he said I should bring the child. When I got there and the SBR was done it was high and above phototherapy level.”. **(FP2, PNW, 2 years).***

Another midwife also mentioned how the sun bathing is a prevailing belief to managing NNJ both by midwives and mothers:

> *“Yes, I have heard some myth from both colleague midwives and parents and some of the myths the parents talked of were that in the olden days if a child is jaundiced the child must be exposed under the sunlight or baby will be given glucose to drink.” **(IDI-P1, NICU, 12 Years).***

A midwife affirms that sunbathing is the most frequent dominant perspectives on NNJ that affect midwives’ response and the management of NNJ is sunbathing:

> *“The misconceptions concerning NNJ are many but the most heard is the exposure of jaundice babies under the sunlight So, for some of them once you mention that the baby is jaundice and has to be admitted they will not agree but there you will hear that a family member’s baby was jaundice and it resolved after exposing the baby under the sunlight and so refuses the admission for the phototherapy to be started.”. **(IDI-P11, NICU, 3 Years).***

According to the midwives below, some mother’s belief the food pregnant women eat causes NNJ which affect midwives’ response to timely management of NNJ:

> *“If you are pregnant and you eat oily food, margarine, fried plantain, oil rice that is what the majority say if you eat that when you are pregnant after delivery your baby will be jaundice”. **(IDI-P7, PNW, 5 years).***

> *“Yes, I have heard mothers saying that when they eat egg the yoke that is yellow during pregnancy, the baby will be jaundice. That is one thing I have heard.” **(IDI-P4, NICU, 1year).***

A participant shared more than one perception of NNJ that affect their care and management of NNJ:

> *“Yes, I have heard several misconceptions. Some of the mothers will not want the admission but will say they will expose the baby under the early morning sunlight (na ewia no behye Jaundice no) and the sunlight will burn the jaundice. Other say if a baby has yellow sclera and you put breastmilk on the eye that too helps. And other also say when you feed babies with colostrum it does not help it causes jaundice.” **(IDI-P6, PNW, 5 Years).***

For the perceptions of NNJ that impacts midwives’ care towards NNJ, according to the midwife below some mothers refuse to feed their babies with colostrum with the belief that it causes NNJ:

> *“Another one that I experience on my ward was a mother whose baby could not feed well on breast and was told to express milk into a cup to feed the child. After expressing the breastmilk, the colostrum that is yellow she said her mother, thus the baby’s grandmother says it will cause the child to be yellow. The other she expressed that was yellow too she did not bring it but threw it away again”. **(FN3, NICU, 3 Years).***

These common perceptions of NNJ shapes midwives care provision and responses towards NNJ. Notwithstanding midwives’ awareness of the condition, they are deeply ingrained with the shared beliefs about NNJ because they are the perceived social behavior towards NNJ in their community.

### 3.23. Neonatal Jaundice Through a Spiritual Lens

This subtheme explained people’s perceptions of neonatal jaundice by associating certain religious and faith-based ideas to NNJ. In the Ghanaian society, a number of childhood illnesses are seen through a spiritual lens and are not thought to be treated in a hospital.

The participant below mentioned some parents associate NNJ to be spiritual and does not warrant to ne managed at the hospital:

> *“I have also heard especially my ward when the mothers come, they say this is not any ailment the green veins on the stomach it is “ASRAM” when I do herbal treatment it will go so the admission will not help the child so discharge my child for me to go and do herbal treatment”. **(FN1, NICU, 3 years).***

Another spiritual aspect of NNJ according to one participant was that some mothers’ belief the condition (NNJ) is bought and given to the babies:

> *“(Eye a omo susu se obi na ato yare3 no ama akora no anaase akora no ye nsuo ba) they belief someone bought and gave the jaundice to the baby or the child is a child from the river gods but studies have made us understand that cerebral palsy and the rest may be a complication of NNJ”. **(IDI-P9, PNW, 5 Years).***

Others exhibit their faith in God and refuse admission when diagnosed this is what a participant mentioned impacted their response to NNJ:

> *“The challenging thing that makes identifying NNJ early on my ward is most of the parents even after identifying and telling them they find it difficult to accept even if she accepts, some of them because of their belief and spirituality they will always say God will do it, God will do it, (“Nyame beye, Nyame beye”). It ever happened on my ward later when the woman reported with the child, she lost the child.” **(FP2, PNW, 2 years).***

The above data indicated that neonatal jaundice is viewed with a spiritual lens in the Ghanaian community and it affects the management of NNJ by midwives.

### 3.24. Following Established NICU Guidelines

The data from the participants indicated that NICU midwives knew and abided by their units’ protocols in identifying and managing NNJ.

A midwife mentioned what she does and detailed the expected protocols to be adhered to on the ward when a baby gets to the NICU with jaundice:

> *“With protocols here we have our protocols, when a baby arrives, we do our assessment, check the vital signs, check the bilirubin with TCB. If they are coming from the OPD they TCB has already been checked for the Dr to diagnose before the baby will report for admission. We take our samples for SBR, the one whose case is severe it is requested as urgent, we check FBC, blood group, G6PD and so when we are done if there is no sepsis apart from the NNJ, some of them when it gets to the extreme that have high temperature so with that if the doctor has requested for antibiotics we start other than that, in the absence of any other diagnosis we start phototherapy and if the baby needs intensive phototherapy with fluids, we set the fluid”. **(IDI-P5, NICU, 5 years).***

Another midwife stated the jaundice babies are admitted and the mothers of the babies are informed:

> *“The guidelines when we identify a jaundice or yellow baby, the first thing is admission, we will inform the mother, take blood samples for laboratory investigations, start phototherapy and make you understand for the day of delivery till the 7 days, your baby will be nurse under the phototherapy, …” **(IDI-P1, NICU, 12 Years).***

A participant from PNW mentioned they do not have any specific guidelines that aligns with their care for babies that develop jaundice:

> *“I don’t think there are specific guidelines but because of the collaboration between NICU and PNW every day, I will have to go through all the mothers so any baby that will be identified to be yellow, whether the doctor has seen or not whether discharged or not, I speak to the mother and send the baby to NICU but I don’t know of any protocol on the ward.”. **(IDI-P6, PNW, 5 years).***

Another PNW midwife also stated they send the babies that are yellow in color to the NICU prior to pediatric review at the PNW:

> *“So, at my unit we don’t wait for the Drs. to come we send those babies to NICU right away even before the Drs. will come so that if there are any laboratory investigations they can take blood samples to know the bilirubin level and start phototherapy early”. **(IDI-P3, PNW, 9 Years).***

Some NICU midwives both from the individual in-depth interviews and also from the focus group discussion also mentioned setting up fluid for babies during intensive phototherapy:

> *“For the children that are severely jaundice we set up fluid, to prevent hypoglycemia and also the frequent the mothers from picking the severely jaundice babies from under the phototherapy for breastfeeding then we manage the child until the child is okay.” **(IDI-P2, NICU, 5 years)***

Some participants also mentioned when the serum blood bilirubin of neonates are exceedingly high exchange blood transfusion are done for those babies:

> *“But when it exceeds a level there is what we call exchange transfusion we will take most of the blood that is affected with bilirubin and give fresh blood without bilirubin that is the main guidelines for those with severe hyperbilirubinemia”. **(IDI-P1, NICU, 12 Years).***

> *“If they are jaundice oh when they report like that, we put them under the phototherapy, some of them too if they are being managed under the phototherapy and the serum bilirubin is very high then there is the need for exchange blood transfusion and it is also done”. **(IDI-P4, NICU 1 Year).***

While majority of the PNW staff were not aware of any protocol at the ward that guides their care for babies that becomes yellow. All the NICU midwives that participated in the study affirmed the availability of a protocol and were committed to adhering to their unit’s protocol.

### 3.3. Social Expectations

These represent the perceived social approval or disapproval of a behavior; these norms reflect what people believe is socially unacceptable or acceptable.

### 3.31. Grandmothers’ and Families’ Disapproval for Hospital Care

In the Ghanaian context care of the baby is a collective care and social influences from family members and grandmothers compelled the mothers to disapprove hospital care.

A participant mentioned notwithstanding the initiation of phototherapy grandmothers will visit and inform the physician to discharge their grandchildren for herbal treatment:

> *“Sometimes after you have started the phototherapy, when the grandmother visits as they are waiting at the waiting area then the grandmother that is the mother of the baby’s mother will be advising the mother so you will see the mother will come for the baby when you inquire then they say the grandmother says it is “ASRAM” inform the doctor to discharge me if I do herbal treatment it will help”. **(FN3, NICU, 3 Years).***

Another participant emphasized how the mothers whose babies are admitted to the NICU for phototherapy listen to their family members more:

> *“…some of them (mothers) listen to their family members more, a grandmother can visit the mother at the mothers’ room after the visit if you don’t check on the mothers may report to the unit with herbal preparation …” **(IDI-P2, NICU, 5 years).***

For a midwife even after the mother of the baby had accepted the admission the grandmother came and disapprove of the grandchild’s admission:

> *“Yes, the mother herself understood but the grandmother they say is the DCE’s wife when she got to the unit, she said she will not understand for the grandchild to be admitted so when I pick the mothers ANC book and asked the mother to follow me with the child to NICU by the time I realized the grandmother had picked the grandchild to their car and left the book with me”. **(IDI-P8, NICU, 9 years).***

Some fathers also refused the admission of their neonates although the mothers may agree for the baby to be admitted on account of neonatal jaundice:

> *“The mother had accepted the admission but the father was refusing the admission. The father too was saying it is his money that will be used for the admission and so will not allow for the admission. He does not have money after paying the wife’s bills, he does not have any other money to pay bills for the baby for the wife to stay back again it a no” **(FN1, NICU, 3 Years).***

### 3.32. Social Approval of Sunbathing

Despite the educational background, exposing infants to the sun has long been accepted as a socially acceptable method of managing NNJ.

A participants affirmed some mothers have accepted sun exposing as the treatment for NNJ, some of her colleague midwives also belief sunbathing is used in managing NNJ:

> *“…the mothers some say the early morning sunlight, when you expose the baby under the sunlight the sunlight will get rid of the bilirubin, some of my colleagues too say that when a baby is exposed under the sun like the mothers”. **(IDI-P5, NICU, 3years).***

> *“Some of the mothers will not want the admission but will say they will expose the baby under the early morning sunlight (na ewia no behye Jaundice no) and the sunlight will burn the jaundice” **(IDI-P6, PNW, 5 years).***

A participant mentioned how a colleague midwife told her to expose the baby under the sun, because she explained it a three-day old baby:

> *“For instance, my own self when I delivered my child it was a Monday and I was supposed to go for postnatal clinic on Wednesday for BCG. When I got there after the nurse held my baby and released her hand that part was yellow and ask me to show it to the midwife. When I got to the midwife, she said three days old baby is physiological and so I should expose her under the early morning sun ….”. **(FP2, PNW, 2 years).***

Exposing babies under the sun due to neonatal jaundice has been a time-honored tradition that dates back to the ancient times:

> *“Yes, I have heard some myth from both colleague midwives and parents and some of the myths the parents talked of were that in the olden days if a child is jaundiced the child must be exposed under the sunlight….” **(IDI-P1, NICU, 12 Years).***

A participant through the interview stated that some mothers refuse admission and by telling the midwives about a family member who exposed a baby under the sun and the NNJ resolved:

> *“The misconceptions concerning NNJ are many but the most heard is the exposure of jaundice babies under the sunlight. So, for some of them once you mention that the baby is jaundice and has to be admitted they will not agree but then you will hear that a family member’s baby was jaundice and it resolved after exposing the baby under the sunlight and so refuses the admission for the phototherapy to be started”. **(IDI-P11, NICU, 3 Years).***

A participant mentioned how cultural practice of sunbathing, in part contributed to infant morbidity:

> *“I encountered a Muslim who has 6 boys all the 6 children are girls the 7^th^ and the only one that was a boy day 2 after delivery the baby was jaundice and she took the baby to a nearby facility (name deidentified), the midwife told the mother it is nothing so she should go home and expose the baby under the sunlight. So, unfortunate the yellow was worsening with days the baby reported to our NICU at age three (3) when the baby was reported to our facility to meet the pediatrician as you mentioned some complications, the child a cannot stand to walk, cannot speak, has cerebral palsy and the child has difficulty hearing”. **(ID1-P9, PNW, 5 Years).***

While mothers and midwives alike knew of family members whose neonates’ NNJ resolved under sun exposure, believing it has roots in tradition. The social approval of sunbathing a baby with jaundice as a treatment for NNJ will affect the midwives’ response to NNJ.

### 3.4. Conformity Pressure

The behaviors that are perceived to be accepted or not accepted socially, and the behaviors within a society, institution, group that are perceived to be prevalent all impact the individual midwife’s behavior and their responses to these norms in the care of the baby. Midwives’ contribution to NNJ Care are influenced by these norms, their understanding of the standard and approved policies in their facility influences them to negotiate care.

### 3.41. Midwives Contribution to Neonatal Jaundice Care

Through their actions, choices, and advocacy, midwives significantly impact the level of care in NNJ. Midwives’ decision-making and behaviors in NNJ care are influenced by their comprehension of best practices, procedures, and rules.

Some participants proactive approaches to detect NNJ and their use of evidence-based practice was evident through the quotes below:

> *“…we do assessment so if through the assessment we detect that a baby is yellow, we use the TCB to check so when we check and see that the bilirubin level is high, we take sample for blood investigation and keep the baby under the phototherapy then we inform Dr. about it”. **(IDI-P5, NICU, 3 years).***

> *“At times when you touch them and release your hands you can see that part is jaundice. Then I will take sample for SBR then start the phototherapy and call the medical officer, to confirm whether the baby needs the phototherapy or not.” **(FN1, NICU, 3years).***

A postnatal midwife mentioned she does not only assess the mothers at the PNW but also observe the babies during her shift:

> *“During handing over I move from bed to bed, so I do not only assess the mothers. I check the babies, their skin color and general appearance”. **(IDI-P9, PNW, 5 Years).***

The application of their midwifery knowledge helps them in identifying and managing NNJ timely to prevent complications of NNJ:

> *“When it is identified early it helps in the prevention of brain disorders because if the baby is managed early with phototherapy, it helps the neonate to get well soon, so I think that if it is identified early and managed promptly the child will be free from neurological disorders as mentioned earlier aha” **(IDI-P7, NICU, 5 years).***

> *“If a child gets jaundiced and is identified early it helps us to transfer the child to where we term NICU for them to start phototherapy, before we start the phototherapy……. Identifying it early prevents it from getting severe and prevents staining of the basal ganglia of the brain with bilirubin” **(IDI-P1, NICU, 12 years).***

The same NICU participant above shared how they initiate treatment without delaying even in the absence of the medical officers:

> *“….. a mother came for third day postnatal clinic and was directed by a midwife to come and see us because the midwife suspected the child was yellow, immediately, I saw the child on the corridor with the midwife you could see from afar that the child was very yellow, we took samples without delaying to the lab and started phototherapy the next day the yellow was reducing…” **(IDI-P1, NICU, 12 years).***

A different NICU midwife also shared what she does in the absence of the medical officers and pediatricians:

> *“…in the absence of the Drs. if a baby is brought to the unit who is jaundice, we first check the bilirubin level with the TCB machine if we see that the bilirubin level is high, we will take sample for serum bilirubin and take G6PD and blood group also and do a laboratory investigation. We will ask the mother to breastfeed the child then take off the neonates clothing then we leave the diaper and then cover the baby’s eyes with an eye shield and we put the child under the phototherapy”. **(IDI-P2, NICU, 5 years).***

### 3.42. Protocol-Informed Shared Decision-Making and Autonomy Balance

The participant replies demonstrate the difficulties in striking a balance between clinical judgment and maternal autonomy. Based on their evaluation of the baby’s condition, some midwives may feel obliged to start phototherapy, while others place more emphasis on cooperation and negotiation with the mother. These characteristics highlight how crucial it is to navigate collaborative decision-making through effective communication, empathy, and negotiation, particularly when there may be divergent opinions on the best course of action.

A participants response address situations where there are discrepancies in situations the care provider’s plan of care is different from the parents. Some participants mentioned how maternal autonomy and medical necessity were balanced:

> *“.. If she still says no and refuses the admission then we are compelled to admit the baby because the baby is a minor and cannot take decision on its own so we will have to take the best decision that will help the child”. **(IDI-P8, PNW, 9 years).***

According to certain participants’ narrations, when it comes to choosing and treating infants with jaundice, medical necessity takes precedence over the mother’s autonomy:

> *“We mostly do what is best for the baby, aha because the child does not only belong to the mother as a minor but also the nation. If even she goes home will come back the following day”. **(FN3, NICU, 3 Years).***

> *“For me for the child to get well is my motive and so I will try to understand whatever any mother does but I will make sure the baby is sent to NICU”. **(IDI-P6, NICU,5 years).***

Some participants have witnessed women refusing admission by exercising their autonomy over the newborn:

> *“So, in our setting the mothers think they have delivered their own children and so have to make most of the decision concerning their children. And we do not have any specific law, if even there is a law it is not strict against them … So, the mothers have effect and influence in the care of their children.” **(IDI-P3, PNW 9 years).***

Another participant discussed how mothers’ experiences affect the choices they make about the health of their infants:

> *“Me too what I know is that, some of them has ever delivered and their babies were jaundice but was not admitted but resolved and nothing happened to them so for those mothers it will take only the last person standing here in the facility to explain things to them. You can tell them everything book and everything home but they will not understand”. **(FP3. PNW, 1 Year).***

> *“Yes, it has happened before the mother said her previous baby was yellow and at the NICU the baby was nursed under blue light and so when the second child was discharged and became yellow on the second day, she did not even bother to bring the baby for admission. She bought a blue fluorescent light and the baby was sleeping under the blue light. By the time she came, complications had already set in and the baby even had an exchange blood transfusion” **(IDI-P10, PNW, 4 years).***

### 3.43. Supervisors’ Expectations: A Normative Influencing on Neonatal Jaundice Care

Concerning the perceptions of anticipated behavior from the socially approved actions, influence emanated from supervisors’ expectations concerning neonatal jaundice care. This highlights how the perceived expectations of authority figures (supervisors) shape the behavior and decision making of midwives in relation to NNJ management.

A participant shared how the supervisors expect that if for nothing at all there should be reduction in the bilirubin levels:

> *“Yes, left to them every 2 days we will have to repeat the SBR, strict monitoring some of them the mother after picking the child to breastfeed it will take quite some time and the child is not put back in the machine our supervisors expect if not for anything there must be a change in the bilirubin level for them to know the bilirubin level is reducing”. **(IDI-P7, NICU, 5 years).***

> *“Our in-charges expect that any baby that we identify to be jaundice we will manage them based on the standard management plan so that the baby can go home well” **(FN2, NICU, 5years).***

Some midwives stressed that their supervisors want them to manage the babies on the ward with jaundice promptly after detecting it:

> *“Yes, please, for expectations they have expectations looking at my unit neonatal jaundice is the highest cause of admission and every year NNJ tops the incidence of conditions on the ward. We get neonatal jaundice and is the majority of cases their expectation is that if for instance is as a result of G6PD is highly unpreventable if the baby becomes jaundice, it is not the fault of the mother and it is also not the fault of the midwives their expectation is for us to provide prompt intervention and prevent complications to our neonates ….” **(IDI-P2, NICU, 5years).***

> *“They have expectations that we can identify it early especially mothers at risk that we may identify them early and start treatment early so that the mother will go happy with the baby”. **(IDI-P6, PNW,5 Years).***

No jaundice identified should become severe, this was what one of the participants said during the interview:

> *“Our supervisors don’t want the situation whereby it (NNJ) will become severe to affect the brains of the neonates we don’t want the case whereby neonatal jaundice will cause neurological disorders at all. So, for the supervisors left to them alone there should always be milder cases that can be manage under the phototherapy for them to go home we don’t want complications of neonatal jaundice.” **(IDI-P3, PNW, 9 Years).***

*Another participant mentioned that their supervisors would want them to inform the appropriate persons if they identify that a newborn is jaundice:*

> *“Please, they expect that we identify it early and after the identification inform the appropriate person for prompt action to be taken”. **(FP2, PNW, 2 Years).***

## Discussion

This study investigated how societal expectations influence midwives’ reactions to neonatal jaundice at Ghana’s St. Patrick’s Hospital in Maase-Offinso. The main conclusions showed how care delivery was impacted by common perceptions regarding NNJ. There is compelling evidence that Ghanaian cultural customs have an impact on children’s health (Nott, 2018). According to the data collected, traditional medical and cultural practices continue to play a major role in the care of babies with jaundice. This demonstrates how, despite civilization, traditional cultural practices continue to play a major role in African health care (Ibrahim & Ahmedolaitan, 2022). Additionally, a study conducted in Ghana demonstrated that community health workers continue to employ home remedies, herbal remedies, and extract from unripe pawpaw water to treat NNJ (Wolski et al., 2024). The shared belief of putting breastmilk on babies eyes and not feeding the newborn with colostrum aligns with the study according to Salia et al., (2021). A different study in Iran also found majority of the mothers will use herbs in managing NNJ (Al-Zamili & Saadoon, 2020). All the above study shows how culture and shared beliefs cannot be separated from health. The studies highlighted above used different but in all of these studies both health care practitioners and mothers held to the shared beliefs.

Mothers declining admission on religious grounds and how NNJ is perceived via a spiritual lens were highlighted. Some participants referred to NNJ as ASRAM, a spiritual condition that complies with (Acheampong et al., 2022). Furthermore, Meñaca et al., (2017) described ASRAM as a spiritual illness that can be transmitted from mother to child.

Mothers’ passions for spiritual care instead of hospital-based treatment were influenced by spiritual beliefs, and they firmly believed nothing will happen to their infants who have jaundice in the name of Jesus. This revelation confirms people’s spirituality and directs their passion and thoughts toward a therapeutic approach (Hvidt et al., 2020). The study’s findings clearly demonstrated the use of herbal remedies, demonstrating how traditional medicine which is frequently firmly anchored in cultural customs offers all-encompassing methods that connect with local populations and attend to both physical and spiritual health issues (Kendi, 2024). According to a recent study conducted in Ghana, religious adults with mental illness may seek mental health care from religious leaders in addition to secular mental health professionals. The study’s conclusion that NNJ is perceived through a spiritual lens and mothers choosing spiritual measures is consistent with this finding. This demonstrates how religion and health-related activities have a complicated relationship (Boateng et al., 2024).

Nearly majority of the participants adhered to the social norm that sunbathing is a long-standing practice that both mothers and midwives consider to be the management of NNJ. One participant suggests exposing infants with jaundice to sunlight, which is consistent with research involving mothers (Huang et al., 2022; Seneadza et al., 2022). One participant described how a mother who had brought her infant to a certain clinic with jaundice was instructed to expose the child to the sun; nevertheless, the mother had reported to her facility after three but the infant had cerebral palsy. This demonstrates indisputably that newborn morbidity and mortality in Ghana have been partially caused by cultural practices and attitudes on child health care (Sumankuuro et al., 2019). This is so because it explains how the practices towards NNJ are driven by deep rooted beliefs and cultural practices other than midwifery practice.

According to the findings, grandmothers disapproved of hospital care since they had more influence and voice over how their grandchildren should be managed than even the child’s parents. This is similar to research by Adama et al. (2021) that discovered that communal living, respect for the elderly can aid in the care of the community’s infants.

According to the research, different midwives prioritize different aspects of shared decision-making, such as clinical judgment, negotiation, and mothers’ autonomy. Midwives made decisions based on their adherence to the ward’s protocol and refraining from behaviors that would violate the ward’s policies. In order to retain their social standing, midwives must make decisions based on empirical expectations, psychological well-being derived from conformity, normative expectations—that is, avoiding activities they believe would be disapproved—and, finally, their personal norms (Gavrilets et al., 2024). It was difficult and stressful to decide whether to admit a newborn with NNJ, especially when the participant thought the parents were adhering to their cultural belief that NNJ was not a serious condition and could be treated at home with sunlight and herbs. This reinforces the claim made by Parish et al. (2022) that families, infants, and medical professionals all naturally experience stress when making decisions. Parents are usually in a better position than others to understand their children’s needs and make intelligent choices about their babies medical treatment because they are mindful of what interest them (Spriggs, 2023). However, the care also took into account the babies’ best interests; according to seven of the participants, if the child needs to be admitted, the parents do not have the last word; instead, what is best for the child is done. This is true according to O’Brien and Newport (2023), some decision-makers were able to find a balance between medical necessity and mother autonomy by using insight, empathy, and a great deal of analytical and communication skills.

In terms of midwives’ contribution to NNJ care, all NICU participants exhibited evidence-based nursing practice. Midwives’ practices significantly shaped NNJ treatment. The regular use of the ward recommendations in the treatment of infants with jaundice may be the cause of this discovery. This result is comparable to a study that shown a favorable substantial impact on either guideline adherence or patient-related nursing outcomes (Spoon et al., 2020).

According to the majority of midwives in this study, a newborn’s jaundice is diagnosed by measuring transcutaneous bilirubin and then confirming the diagnosis with a blood sample for total serum bilirubin following visual inspection. This is consistent with the findings of Nambinga and Nghitanwa, (2023) study. The results may be congruent probably because the midwives used for the study confirmed it is the top cause of admission at the unit and so they have been caring and managing for NNJ cases and so know and are equipped in managing the condition they daily manage.

### Strength

The study strengths include its contextual relevance, in-depth understanding of established convention, identification of social expectations, identification of key influencers (colleagues, supervisors and grandmothers) who shape midwives’ practices, informing targeted interventions and contribution to knowledge on NNJ care in Ghana.

### Limitations

The first limitation of the study is its geographic location. Because of cultural differences, the results might not apply to other areas. The accuracy of the transcription and translation may have been impacted, potentially affecting the interpretation of the results, since data collection was done in Ashanti Twi and participant codes were switched to English. Parents’ opinions on the early detection and treatment of NNJ were not included in the study, which only examined the experiences of midwives.

## Conclusion

The study explored how social expectations shape midwives’ management of neonatal jaundice at St Patrick’s Hospital in Maase-Offinso, Ghana, focusing on the interplay between cultural beliefs, community expectations, and clinical practice. The study revealed that shared beliefs, spiritual beliefs about NNJ in the Ghanaian community negatively impacted midwives’ responses to care. Social approval of sunbathing and grandmothers’ disapproval of hospital care highlighted the influence of deep-rooted cultural practices on NNJ management, often superseding midwifery expertise. The study also highlighted challenges in balancing clinical judgment with maternal autonomy, particularly when grandmothers’ decisions were respected. Conversely, midwives’ professional norms and supervisors’ expectations positively influenced NNJ care provision, underscoring the importance of leveraging social norms to improve care outcomes.

## Data Availability

Anonymized data excerpts are included in the article.

## Notes

### Competing Interest Statement

The authors have declared no competing interest.

### Funding Statement

The author(s) received no specific funding for this work.

### Author Declarations

Christian Health Association of Ghana (CHAG) Research Department - Institutional Review Board (IRB). Written consent was used for this stidy, the approval ID for the study is CHAG-IRB05042024. https://sites.google.com/view/chag-research/institutional-review-board

